# OptiLITT: A Computer-Assisted Planning System for Dual-Fiber Laser Interstitial Thermal Therapy using Cylindrical Ablation Optimization

**DOI:** 10.64898/2026.07.03.26356873

**Authors:** Natalie Yeung, Akash Mishra, Ashesh D. Mehta

## Abstract

Laser Interstitial Thermal Therapy (LITT) is a minimally invasive neurosurgical technique in which a stereotactically-implanted fiber delivers thermal energy to ablate intracranial lesions. Existing computer-assisted planning systems optimize trajectories against a one-dimensional line abstraction, then approximate the ablation zone as a fixed-radius cylinder post-hoc to estimate coverage. Trajectories selected as optimal under this model are not guaranteed to remain optimal once the cylindrical extent is applied, which introduces a mismatch between predicted and true ablation coverage. This also may underestimate spillover into surrounding healthy tissue. We present `OptiLITT`, a treatment planning system that represents the laser probe as a cylindrical ablation volume from the onset of optimization, jointly solving dual-fiber placement, lesion coverage, and healthy-tissue spillover as a single coupled problem. All planning parameters are exposed through a user-configurable graphical user interface supporting intraoperative refinement between planning stages.

## 1 Introduction

### 1.1 Clinical Motivation

Laser interstitial thermal therapy (LITT) is a neurosurgical approach in which the surgeon can access and treat pathologic regions and processes that are deep within the brain using small, sub-centimeter burr holes. The therapy may be visualized in real time via magnetic resonance (MR) thermography. This technique can be used to treat a range of epileptogenic foci and brain tumors [1]. A key benefit of LITT, compared to conventional neurosurgical techniques such as open craniotomy, is that patients experience earlier hospital discharge, fewer adverse effects, and decreased use of postoperative opioid-based pain medications [2, 3].

Conventional surgical planning and workflow for LITT relies on readily-available trajectory planning software that are optimized for biopsies and stereoelectroencephalography (SEEG) electrode implantation [4]. More recently, these algorithms have incorporated atlas-based parcellations to constrain trajectories to specific cortical and white matter regions and to avoid critical structures such as vasculature and ventricles. While this approach is effective for simple spherical targets, line-based optimization fails for irregular or elongated geometries where the volumetric extent of the ablation zone cannot be accurately captured by a one-dimensional centerline model. Furthermore, the majority of current approaches do not support deployment of multiple therapeutic devices to the target volume simultaneously. This limitation is clinically significant, as non-spherical structures, including the hippocampus or corpus callosum, may require alternative approaches for optimal ablation [5].

Here, we introduce `OptiLITT,` a surgical planning system for rapid dual-fiber trajectory optimization built around a cylindrical ablation model. All planning parameters are presented via a user-configurable graphical user interface supporting intraoperative refinement between planning stages. We demonstrate end-to-end system application through a proof-of-concept case study, and benchmark the differential-evolution optimizer against four gradient-free search strategies.

### 1.2 Related Work

Computer-assisted LITT planning has been approached along three main directions. The first, and most clinically mature, consist of atlas- and rule-based planners such as EpiNav [6]. These systems rely on whole-brain atlas parcellations to define candidate entry regions, target translations, and erosion depths. Each combination is scored by a cumulative risk function evaluated at 128 equally spaced nodes along the fiber, where the risk at each node reflects its distance to the nearest critical structure. The ablation zone is identified using a line-to-point proximity test against a single fixed-diameter cylinder, reducing coverage estimation to a one-dimensional check rather than a volumetric intersection. This paradigm carries two structural limitations that *OptiLITT* is designed to address. First, dependence on anatomical or functional atlases may introduce segmentation error into the plan, which manifests as overestimation of hippocampal volume in mesial temporal sclerosis or degraded performance on pediatric cortical morphology [7]. Second, this approach is largely constrained to the amygdalohippocampal complex, whereas the application of LITT may extend to other subcortical or cortical lesion locations. *OptiLITT* addresses these limitations by replacing atlas dependence with direct spline-interpolated lesion annotation to support arbitrary lesion locations and types.

A second line of work comprises machine-learning-based planners. A leading LITT-specific example is Li et al. [8], where the study team trained random-forest and linear-regression models on EpiNav-derived candidate trajectories to predict a composite ablation score that balanced amygdalohippocampal coverage against parahippocampal damage. Each candidate trajectory was defined by three discretized design variables (entry gyrus, target translation, and erosion depth). This approach served as a fast surrogate for EpiNav’s exhaustive search rather than a trajectory planner. Since the model is trained on population-averaged trajectories rather than individual patient anatomy, the parameters it identifies as optimal may not generalize to individual patients. More recent studies have focused on applying reinforcement learning to general neurosurgical path planning. For example, Dundar et al. [9] applied Q-learning, in which an agent incrementally builds a path step by step, earning reward for progress toward the tumor and penalty for intersecting critical structures such as vessels, dural sinuses, and white-matter tracts. This produces nonlinear paths suited to corridors that require curvature. LITT trajectories require no such construction, since the applicator is a rigid fiber advanced along a single straight stereotactic line between its entry and target points. This reduces LITT trajectory planning to a direct search over these two points, rather than the sequential decision process that reinforcement learning is optimized for.

A third line of work formulates the trajectory selection problem as a sampling-based geometric search rather than ranking or learning. Zelmann et al. [10] studied how SEEG electrode placement can be optimized by drawing thousands of candidate entry and target points via Gaussian sampling, scoring them on each contact’s recording strength and surgical risk (insertion angle, critical-structure proximity, etc.), and selecting an electrode set from this pool based on these two measures. *OptiLITT* shares this approach’s population-level search, but changes what the objective rewards and how the instrument itself is represented. Each SEEG electrode is a thin shaft with discrete point contacts, which Zelmann et al. model as a line, with each contact’s recording strength decaying as the inverse square of distance. Thermal ablation from LITT produces a fixed-radius zone of tissue destruction, hence allowing *OptiLITT* to model each laser applicator as a finite-radius cylinder rasterized against a millimeter-scale lesion-occupancy grid. The objective accordingly scores both lesion coverage and healthy-tissue spillover, a criterion that a recording-maximization objective cannot capture. Coverage, spillover, and inter-applicator overlap are scored jointly for both applicators within a single objective (Section 3.4), rather than scored per-trajectory and combined combinatorially as in Zelmann et al.

## 2 System Interface and Clinical Workflow

*OptiLITT* implements a six-stage interactive pipeline illustrated in Figure 1. The system automates the computationally intensive stages of LITT treatment planning, including skull surface extraction, lesion volume reconstruction, and dual-trajectory optimization, while preserving manual control over the steps that require clinical judgment: lesion boundary annotation, mid-sagittal plane selection, anterior–posterior commissure identification, and entry-region definition. The system is packaged as a self-contained executable for Windows, macOS, and Linux.

**Figure 1:**
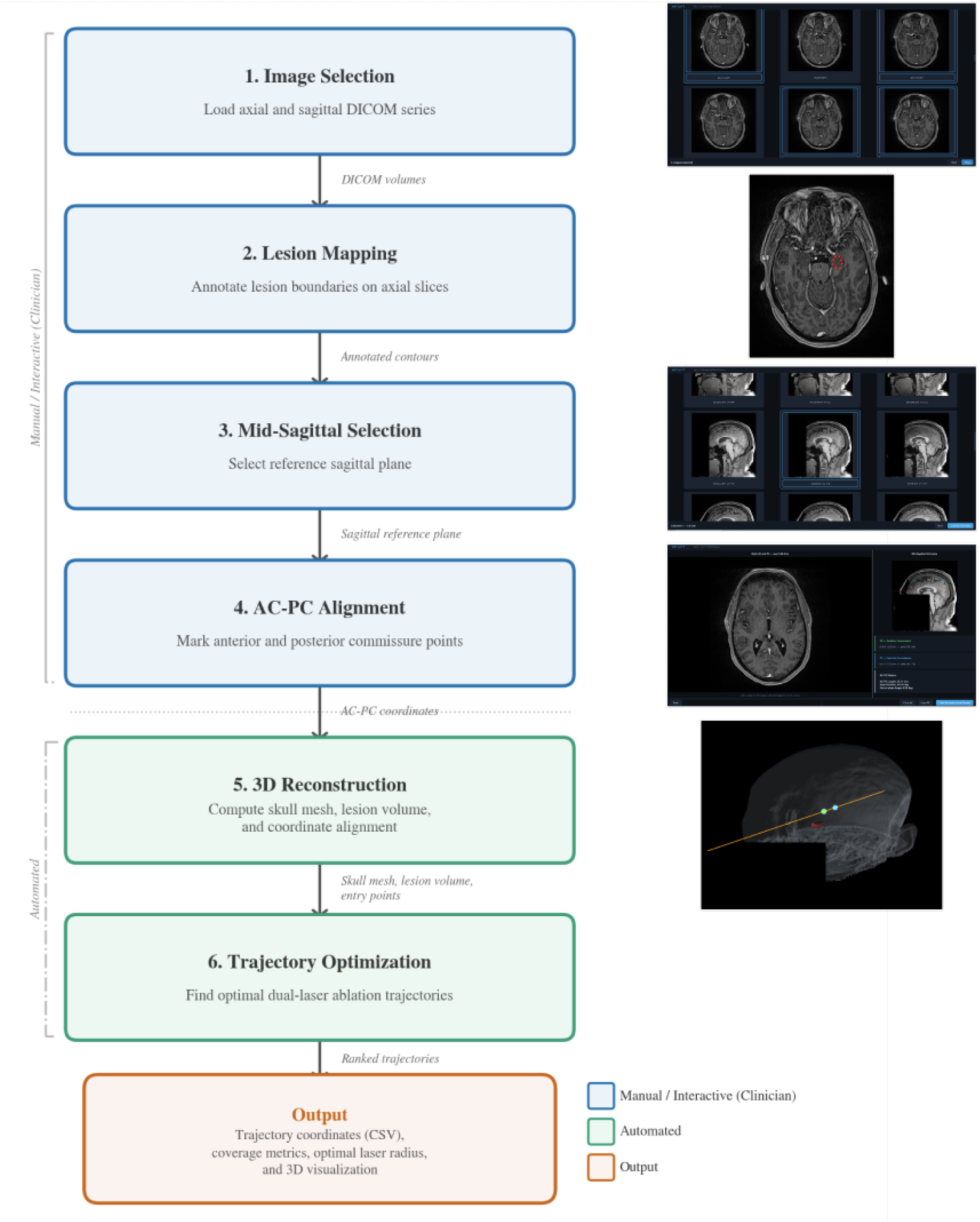
Flowchart of the OptiLITT GUI.

### 2.1 Image Requirements and Outputs

*OptiLITT* accepts any directory containing DICOM files as input and requires both an axial and a sagittal T1-weighted contrast-enhanced MRI series. Since clinical scanners typically acquire images in a single orientation per sequence, the user may re-slice the acquired volume into the complementary orientation prior to loading. This can be performed using tools such as 3D Slicer, ITK-SNAP, or OsiriX, or through the scanner manufacturer’s post-processing console. Files are validated via header parsing and spatially sorted by the `ImagePositionPatient` tag. Oblique scan orientations are handled by extracting direction cosines from the `ImageOrientationPatient` tag, enabling accurate cross-plane coordinate mapping between the two series. Pixel arrays are normalized to an 8-bit intensity range regardless of source bit depth, ensuring consistent display across scanner vendors and acquisition protocols. Upon completion, *OptiLITT* exports a CSV file in which each row corresponds to a ranked trajectory pair in patient (DICOM) coordinate space (mm). This can be directly imported into Python or MATLAB for cohort-level analysis.

### 2.2 Lesion Mapping

*OptiLITT* renders all slices from the user-provided axial directory into a scrollable grid sorted by increasing z-position, from which the user selects the subset containing the lesion. Each selected slice is displayed individually for annotation, where the user clicks a minimum of five points along the lesion boundary. A greater number of boundary points yields a more precise contour, particularly along sharply curved or irregular boundaries.

### 2.3 Anatomical Reference Alignment

To support valid entry region identification, *OptiLITT* provides an interface for manual identification of the mid-sagittal plane and the anterior–posterior commissure (AC–PC) axis. The mid-sagittal plane restricts candidate entry points to the hemisphere containing the lesion, while the AC–PC axis provides a standard anatomical reference overlaid on the skull reconstruction during trajectory review. For AC–PC identification, the system presents axial slices from the 33rd to 67th percentile of the axial stack alongside the chosen mid-sagittal image. The user may mark both commissures on a single slice or across two slices, with pixel positions converted to three-dimensional millimetre coordinates using DICOM pixel spacing and slice position metadata. The system then computes the AC–PC vector and reports the commissural distance, in-plane tilt, and out-of-plane angle. The resulting axis is projected onto the sagittal image for visual confirmation and rendered as a three-dimensional overlay on the skull surface throughout the remainder of the workflow.

### 2.4 Geometric Reconstruction

Following image loading and lesion annotation, the system automatically reconstructs two three-dimensional surfaces (Stage 5, Figure 1): a triangulated mesh of the outer skull surface and a volumetric lesion representation used as the optimization target. The resulting surfaces are rendered in an interactive 3D viewer, providing a spatially accurate confirmation of lesion position, geometry, and AC–PC alignment prior to trajectory optimization.

#### 2.4.1 Skull Surface Reconstruction

Skull surface reconstruction is performed directly from the loaded MRI volume without requiring a separate CT acquisition or segmentation atlas, as the skull model serves solely to define the outer surface geometry for entry point sampling. Intensity normalization uses a single global Otsu threshold across all slices rather than per-slice, avoiding boundary discontinuities at slice transitions. Each slice is binarized against this threshold and the skull boundary is isolated as the outermost voxel layer of the morphologically filled region. Overlapping acquisitions are resolved by grouping and averaging slices within a 0.01 mm z-tolerance, and volumes with non-uniform z-spacing are resampled onto a uniform grid by trilinear interpolation prior to surface extraction. The boundary volume is smoothed with an anisotropic Gaussian kernel to account for voxel spacing differences between in-plane and through-plane dimensions. A triangle mesh is then extracted via marching cubes at the boundary between skull and non-skull voxels, producing a smooth closed surface suitable for 3D rendering and entry point sampling.

#### 2.4.2 Lesion Volume Reconstruction

The lesion volume is reconstructed from the boundary points placed during the lesion annotation stage (Section 2.2). All annotated points on each slice are fitted with a closed spline to trace the lesion boundary regardless of their original placement order. Since users may annotate only a subset of slices containing the lesion, cubic spline interpolation is applied along the z-axis between consecutive annotated contours to fill intermediate gaps. Each interpolated contour is then replicated across all slices sharing that z-position to account for the finite physical thickness of each DICOM slice. The resulting contours are rasterized into binary masks and assembled into a three-dimensional binary volume.

### 2.5 Generating the Valid Entry Region

The valid entry region is defined interactively on the three-dimensional skull reconstruction through an OpenGL viewport (Figure 2). User clicks are unprojected from screen coordinates into scene space and snapped to the nearest front-facing skull vertex, ensuring every placed point lies exactly on the reconstructed outer surface regardless of viewing angle. The region is formed by the intersection of two boundaries on the skull surface. The first (the exclusion zone for face areas) is a horizontal boundary established by clicking points along the eyebrow line. A closed periodic spline is fitted through the mean height of these points, and all skull vertices below this threshold are excluded. The second (exclusion zone for cross-hemispheric trajectories) is a coronal boundary drawn across the superior surface of the skull. The first click is automatically snapped to the horizontal spline to connect the two boundaries, and an open spline is fitted through the remaining points in click order. A hemisphere filter derived from the mid-sagittal plane established during AC–PC alignment (Section 2.3) further restricts candidates to the skull surface on the same side as the lesion. If the resulting set of entry points exceeds the configured maximum, farthest-point sampling selects the most spatially distributed subset, yielding up to 500 entry points passed to the trajectory optimizer.

**Figure 2:**
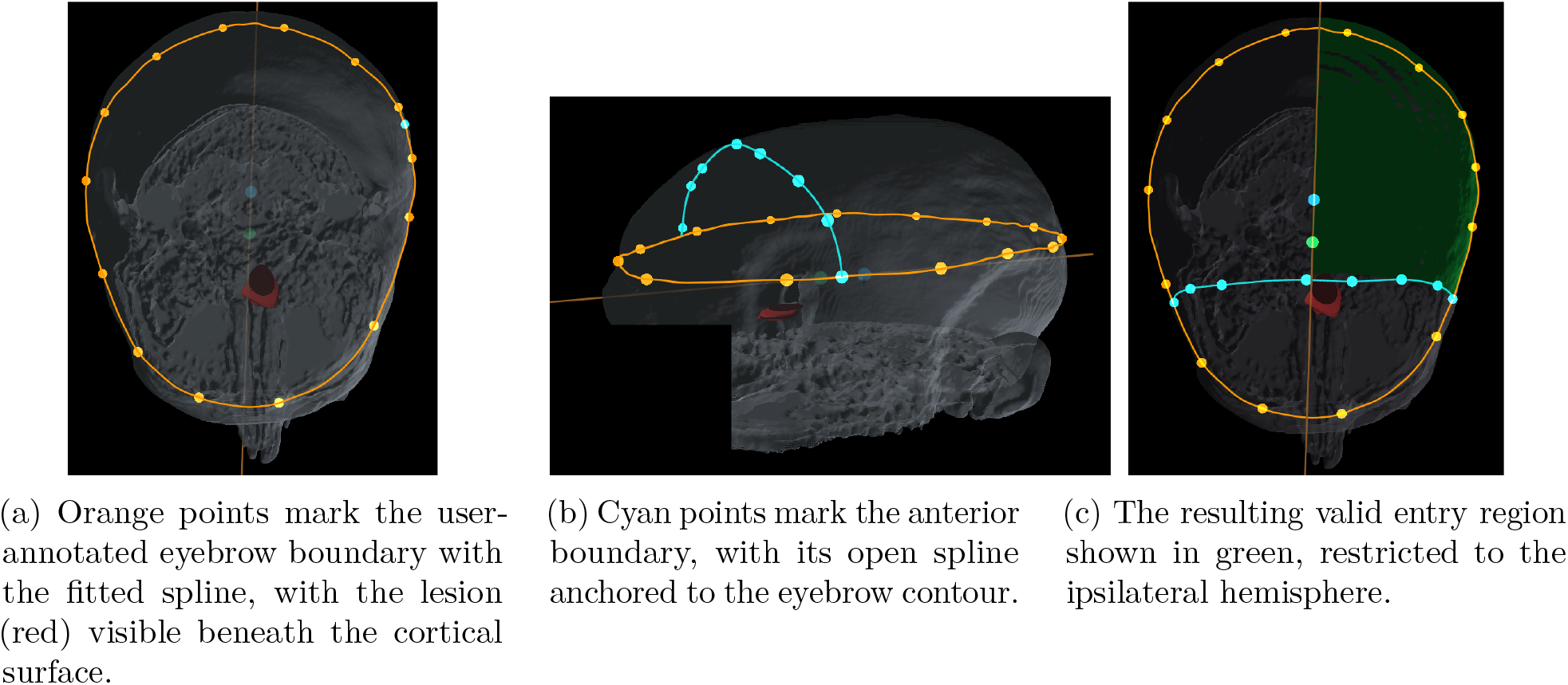
GUI panel for manual valid entry region definition.

## 3 Trajectory Optimization

### 3.1 Cylindrical 3D Volume Optimization

*OptiLITT* replaces the conventional line abstraction with an explicit cylindrical volume model where each laser trajectory *γ* extends from the skull entry point to the far intersection with the lesion bounding sphere. Whereas line-based planning treats the applicator as a zero-width segment, the ablation zone is modeled as a swept cylinder of effective thermal ablation radius *r*, comprising all points whose perpendicular distance to *γ* does not exceed *r*. The lesion point cloud is rasterized into a voxel occupancy grid with a 1 mm cell size. Each occupied cell is dilated by one voxel so that samples within 1 mm of the lesion boundary are classified as lesion interior, enabling O(1) spatial lookups during objective evaluation regardless of lesion size. Lesion coverage is defined as the fraction of lesion volume enclosed by the cylinder

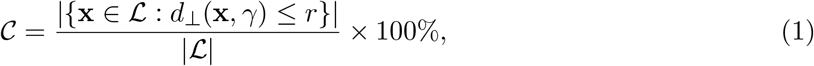

where ℒ is the set of lesion voxels in physical coordinates and *d*⊥ (**x**, *γ*) is the perpendicular distance from voxel **x** to *γ*. Spillover is evaluated over the same grid by sampling the cylinder’s interior directly. At each point along the centerline, 16 cross-sectional samples are drawn via equal-area disk quadrature and classified as lesion interior or healthy, so the spillover penalty reflects the fraction of the cylinder’s continuous volume extending into healthy tissue. For dual-trajectory configurations, both coverage and spillover are computed over the union of both ablation zones.

### 3.2 Poincaré Disk Representation

This section defines the geometric basis shared by the trajectory optimizer and the morphology classifier (Section 3.3), namely a projection of the lesion onto the Poincaré disk ⅅ = {*z* ∈ ℂ:|<> 1}. Each lesion point *p*_*k*_ is taken relative to the centroid 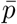 and expressed in spherical coordinates, with radius 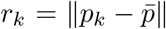, azimuth 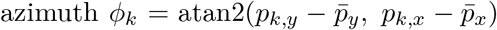, and elevation 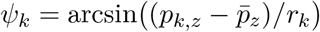. Each point is then mapped to

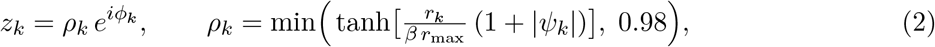

where *r*_max_ = max_*k*_ *r*_*k*_ and *β* = 1.5. The lesion centroid maps to the origin of ⅅ and peripheral lesion points toward its boundary. Since *r*_*k*_ appears only through the ratio *r*_*k*_*/r*_max_, the tanh compression produces a size-invariant measure of peripherality that carries the same geometric meaning regardless of absolute lesion size. The elevation factor (1 + *ψ*_*k*_) augments the projected radius of out-of-plane points, preserving elevation information within the two-dimensional projection.

The Poincaré disk is chosen over a plain polar or stereographic projection because the hyperbolic metric

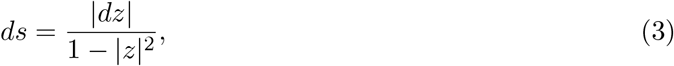

diverges as |*z*| → 1, so trajectories passing through the lesion periphery, which project near the disk boundary, accumulate arbitrarily large arc length. The discrete hyperbolic arc length of a projected trajectory *γ*_*i*_ under this metric is given by

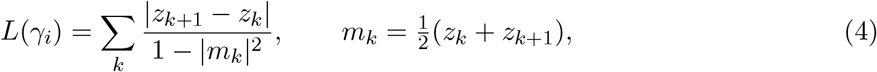

where *m*_*k*_ is the midpoint of each projected segment. This property is later exploited by the morphology classifier (Section 3.3) and the centrality penalty (Section 3.4).

### 3.3 Morphology-Aware Optimization

Prior to optimization, lesion morphology is classified automatically by combining a three-dimensional PCA pre-check with angular and radial analysis on the Poincaré disk of Section 3.2 (Figure 3). The classifier returns one of three morphological classes—spherical, C-shape, or irregular— and is preceded by a post-classifier override (detailed below) that takes precedence over all three tests when disjoint lesion regions are annotated. The classification sequence is outlined in Algorithm 1.

**Figure 3:**
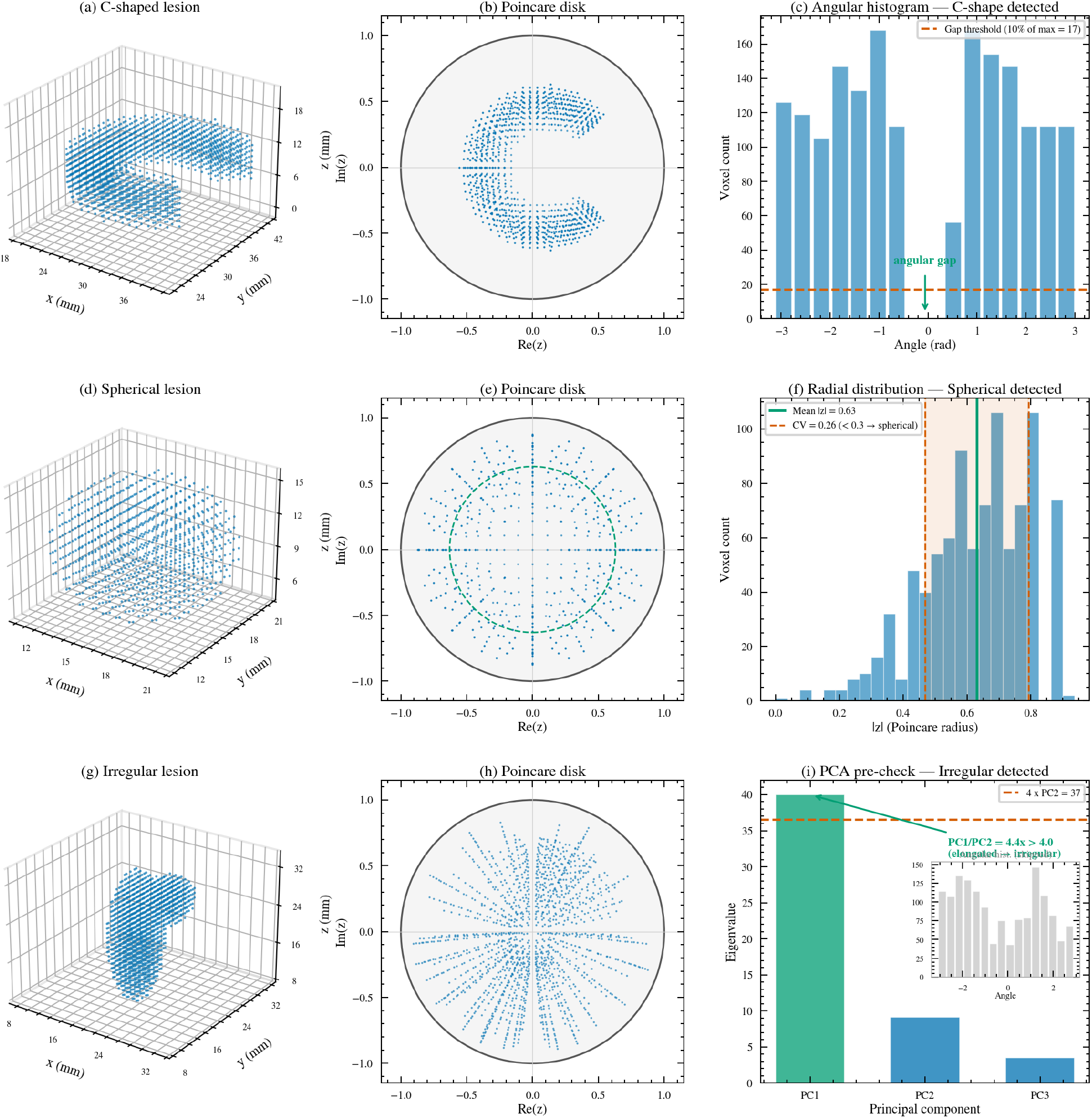
(a–c) C-shape lesion. The angular gap in the torus maps to a corresponding void on the Poincaré disk (b). Histogram bins below 10% of the maximum (dashed line) detect the missing wedge, triggering C-shape classification (c). (d–f) Spherical lesion. The disk projection is radially uniform (e), with the dashed ring at the mean radius. A coefficient of variation of 0.26, below the 0.3 threshold, confirms spherical classification (f). (g–i) Irregular lesion with an asymmetric bulge. The PCA eigenvalue pre-check catches the elongation (PC1/PC2 = 4.8 *>* 4.0 threshold) before the angular histogram is consulted (i.e. the grayed inset).

#### Algorithm 1

Morphology Classification

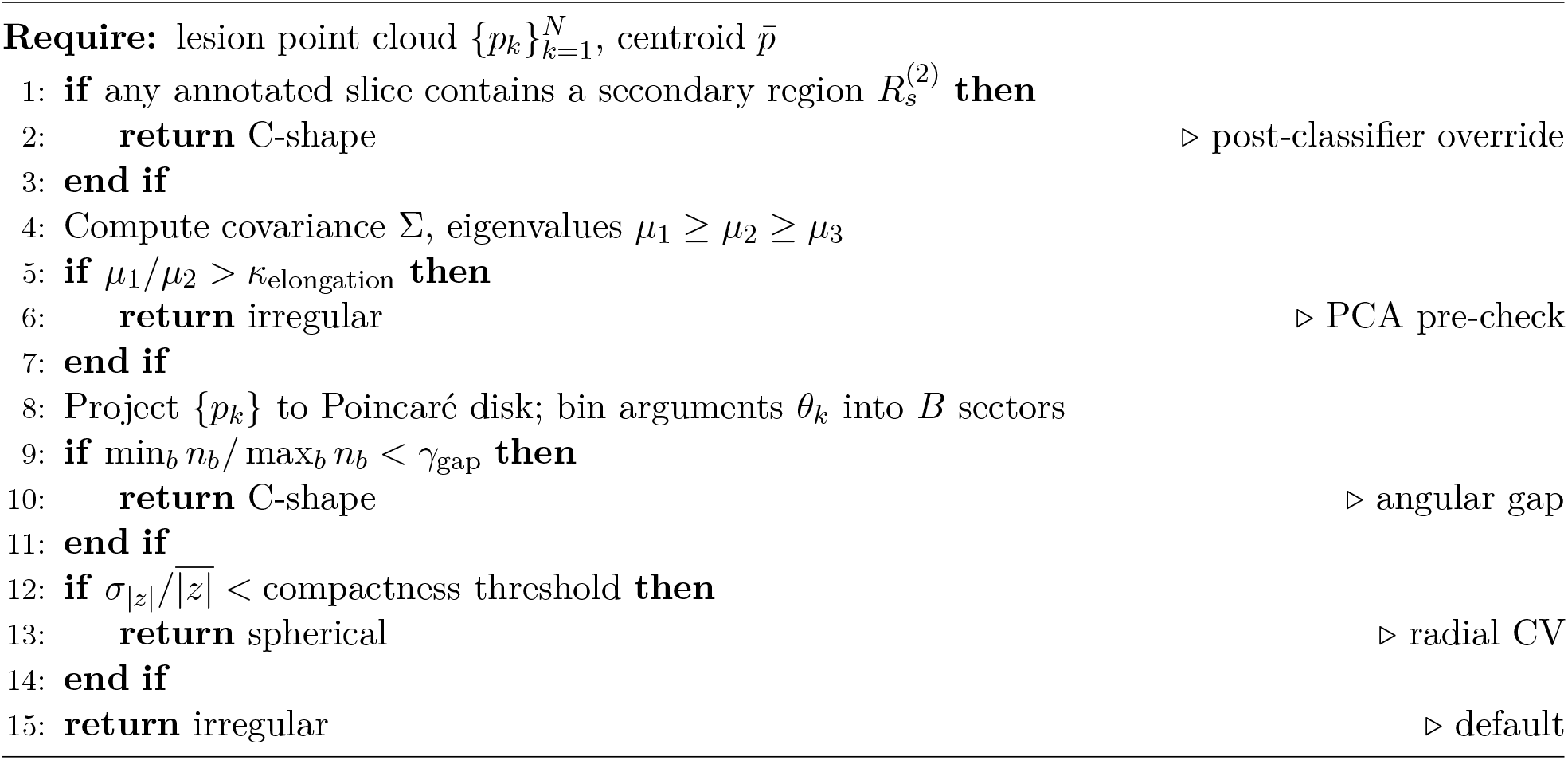

Given the lesion point cloud 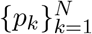 with centroid *p̄*, the covariance matrix

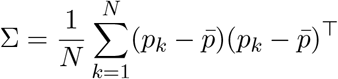

yields ordered eigenvalues *µ*_1_ *µ*_2_ *µ*_3_. The PCA pre-check classifies a lesion as irregular when *µ*_1_*/µ*_2_ *> κ*_elongation_. This test precedes the disk projection because an elongated lesion collapses onto a few dominant angular directions, leaving the remaining sectors empty and causing it to be misclassified as a C-shape if applied after projection. For lesions that do not exceed the elongation threshold, the angular coordinates *θ*_*k*_ = arg(*z*_*k*_) of the disk-projected points (Eq. (2)) are binned into *B* equally spaced sectors. A value of min_*b*_ *n*_*b*_*/* max_*b*_ *n*_*b*_ below *γ*_gap_ indicates a continuous gap in the angular distribution, characteristic of a crescent or toroidal lesion, and triggers C-shape classification. Lesions passing both tests are classified as spherical when the radial coefficient of variation 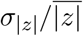 on the disk falls below a compactness threshold, and default to irregular otherwise. The classified morphology directly determines the optimization approach in two ways. First, the target point parameterization is adapted per class. Spherical lesions offset each target along the entry-to-center direction by up to half the lesion radius. C-shape lesions anchor each target at its section centroid with a bounded ± *π/*4 angular perturbation about the lesion center, preventing the optimizer from collapsing a laser onto a thin arc edge. Irregular lesions place targets within a PCA-aligned ellipsoid scaled to half the principal semi-axes, as the geometry imposes no natural placement axis. Second, the inter-fiber overlap and hyperbolic centrality penalty weights, which appear in the objective (Section 3.4), are tuned per morphology class, while spillover and entry-to-center distance penalties are shared across all classes. Spherical lesions additionally receive a perpendicularity bonus. Each candidate solution is encoded as a normalized vector in [0, 1]^*n*^, where *n* = 4 for spherical and C-shape lesions (one entry-point index and one scalar target parameter per laser) and *n* = 6 for irregular lesions (one entry-point index and one azimuth-elevation pair per laser).

The classifier assumes a single connected point cloud, an assumption that breaks down when multiple disjoint regions are annotated. When two disjoint contours appear on the same axial slice, vertical interpolation produces a topologically connected mesh containing an interior void between the two arcs, collapsing the concavity that defines the C-shape and driving the angular gap on the Poincaré disk below *γ*_gap_. The geometry is consequently misrouted to the irregular branch despite its fundamentally C-shape character. The C-shape branch naturally accommodates this structure by partitioning the lesion into two angular sections and assigning one laser per section: after stacking, each user-marked contour forms one angular section with a low-density angle persisting between them, and the partitioner identifies this inter-arc gap to split the point cloud, directing each laser toward its own arc. The post-classifier override (Algorithm 1, line 1) corrects this misrouting, with all downstream optimization parameters drawn directly from the standard C-shape branch.

### 3.4 Dual-trajectory Co-Optimization

Unlike sequential single-fiber planning where the second fiber is fitted conditional on the first, *OptiLITT* encodes the entry point and intracranial target of both applicators in a single parameter vector, so every candidate evaluated during the search constitutes a complete two-fiber plan. For complex morphologies such as C-shape or multi-lobed lesions, this co-optimization implicitly assigns each fiber to a distinct anatomical sub-region of the target without any manual ROI segmentation.

The composite objective scored for every candidate plan is given by

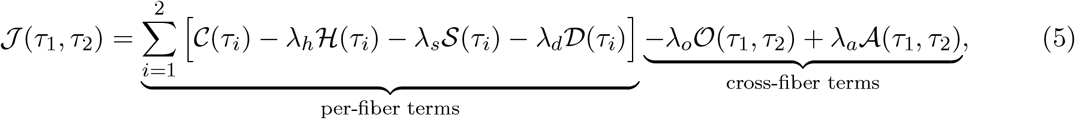

which the optimizer maximizes over both fibers jointly (equivalently, the search minimizes − *J*). The per-fiber terms quantify how each fiber’s cylindrical ablation zone covers the lesion (*C*, Eq. (1)), spills beyond it into healthy tissue (*S*), penalizes deviation from the lesion center via the hyperbolic arc length *H*(*τ*_*i*_) = *L*(*γ*_*i*_) (Eq. (4)), and minimizes the intracranial distance from the skull entry point to the target (*D*). Joint optimization is instead enforced by the cross-fiber terms: *O* penalizes redundant ablation where the two zones overlap, guiding the search toward complementary coverage of the lesion, and *A* rewards orthogonal fiber orientations for spherical lesion geometries. For non-spherical or multi-lobed morphologies is *A* inactive, and complementary placement emerges from the coverage, spillover, and overlap terms alone. Both cross-fiber terms are symmetric (*J*(*τ*_1_, *τ*_2_) = *J*(*τ*_2_, *τ*_1_)), hence the two fibers of any *OptiLITT* trajectory pair are interchangeable. The penalty weights *λ*_*h*_, *λ*_*s*_, *λ*_*d*_, *λ*_*o*_, and *λ*_*a*_ are configured per morphology class (Section 3.3); their values, along with the explicit forms of *C, S, D, O*, and *A*, are protected under a provisional patent application and are not disclosed here.

### 3.5 Differential Evolution Search

Trajectory optimization is formulated as a mixed discrete-continuous problem solved by differential evolution (DE), a population-based global optimizer that iteratively refines a fixed-size set of candidate solutions via mutation, crossover, and greedy selection [11]. Each candidate solution encodes a discrete entry-point selection and a class-specific target parameterization for both lasers simultaneously, with parameter dimensionality varying by lesion class as detailed in Section 3.3. A population of *P* candidates is evolved over *G* generations, giving a total evaluation budget of *P*×*G* objective evaluations. Upon completion, the top-ranked candidates are re-evaluated at full spatial resolution and filtered for diversity using a combined spatial-angular distance metric, ensuring that the final trajectory pairs presented to the user represent genuinely distinct insertion plans rather than minor perturbations of a single solution.

### 3.6 Lesion-Adaptive Laser Radius Selection

In conventional LITT planning, trajectory optimization and ablation radius selection are treated as separate steps, with the trajectory first planned using neuronavigation software and the radius estimated post-hoc from manufacturer data and clinical experience. This leaves surgeons with no systematic means of exploring the coverage–selectivity tradeoff prior to committing to a plan.

*OptiLITT* decouples trajectory optimization from radius selection through a two-stage planning framework. In the first stage, the optimizer identifies the optimal dual-trajectory configuration using an initialization radius scaled to 50% of the patient-specific lesion bounding radius, giving both fibers comparable coverage contributions at initialization rather than letting one dominate from the outset. In the second stage, a binary search over physically achievable ablation radii evaluates coverage along the fixed trajectories, identifying two clinically significant bounds: a lower bound below which lesion coverage becomes inadequate, and an upper bound beyond which lesion coverage plateaus and spillover increases. Both thresholds are configurable at runtime and default to 85% and 95% of total lesion coverage, corresponding to the clinically desired range of ablation completeness. The planner also exposes a fixed-radius override that disables this dynamic sweep, allowing a device-specific or clinically motivated radius to be applied directly for both optimization and coverage evaluation.

## 4 Results and Discussion

### 4.1 Retrospective Application

We retrospectively applied `OptiLITT` to imaging data from one patient with right mesial temporal lobe epilepsy who was treated at North Shore University Hospital, Manhasset, NY (Figure 4a–b). Axial and sagittal gadolinium-enhanced T1-weighted (T1w; MPRAGE) MRI sequences were used as input, acquired at 3 T with 1 mm slice thickness and sub-millimeter in-plane resolution (0.5 mm axial, 0.7 mm sagittal). Both *OptiLITT* ‘s cylindrical volume optimization and a line-based baseline were applied to the same patient imaging to compare lesion coverage and healthy-tissue spillover under identical entry-point constraints and lesion classification.

**Figure 4:**
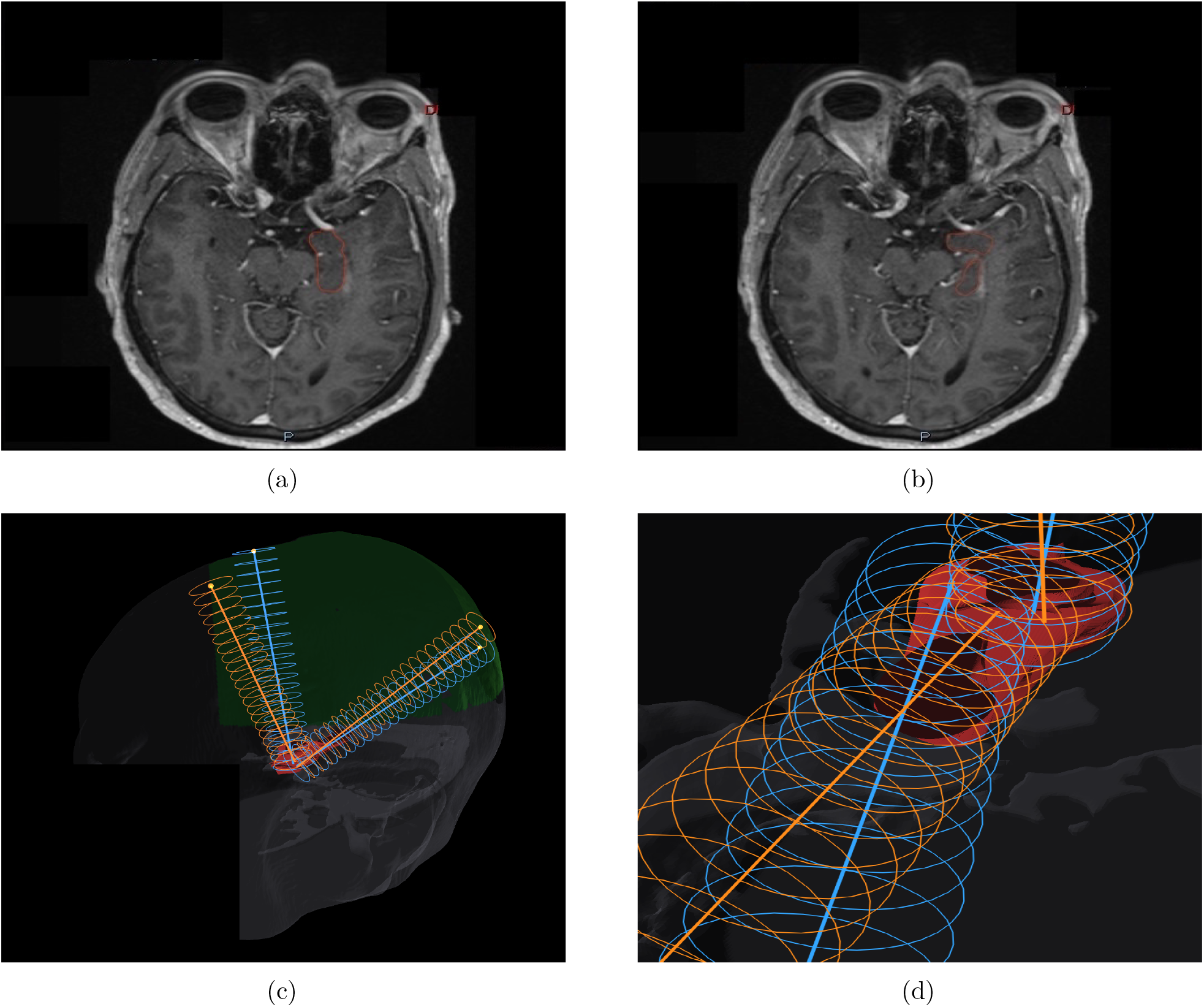
(a) Representative axial MRI slice from one patient showing a fully connected lesion annotation, typical of spherical or standard C-shape classification. (b) A second representative slice showing two discrete lesion regions separated by a healthy intra-void, invoking *OptiLITT* ‘s C-shape post-classifier override. (c) Overview of *OptiLITT* ‘s cylindrical volume-optimized dual trajectories (orange) compared to line-based optimization (blue) overlaid on the reconstructed lesion mesh. (d) Zoomed view showing the trajectory centerlines alongside healthy-tissue spillover from the line-based ablation zone and lesion voxels missed by the line model but recovered by *OptiLITT* ‘s cylindrical ablation model.

The line-based comparison reused the *OptiLITT* optimization framework with one modification. The cylindrical coverage term was replaced by a centerline-proximity criterion minimizing the mean lesion-voxel-to-centerline distance. This draws each trajectory toward the densest part of its assigned lesion region rather than its strict geometric center. The resulting centerlines were dilated to 10 mm, matching the ablation radius used in both configurations.

The *OptiLITT* -planned configuration achieved 99.29% lesion coverage, compared to 97.31% for the line-based baseline. While the coverage difference appears modest, the line-based approach ablated 11.7% more healthy tissue (as a fraction of intracranial volume) than `OptiLITT` at the top-ranked configuration. The geometric basis for this difference is illustrated in Figure 4c–d, and Figure 5 further contextualizes this trend across all candidate configurations, showing that *OptiLITT* trajectories consistently achieve higher lesion coverage with less healthy tissue ablated.

**Figure 5:**
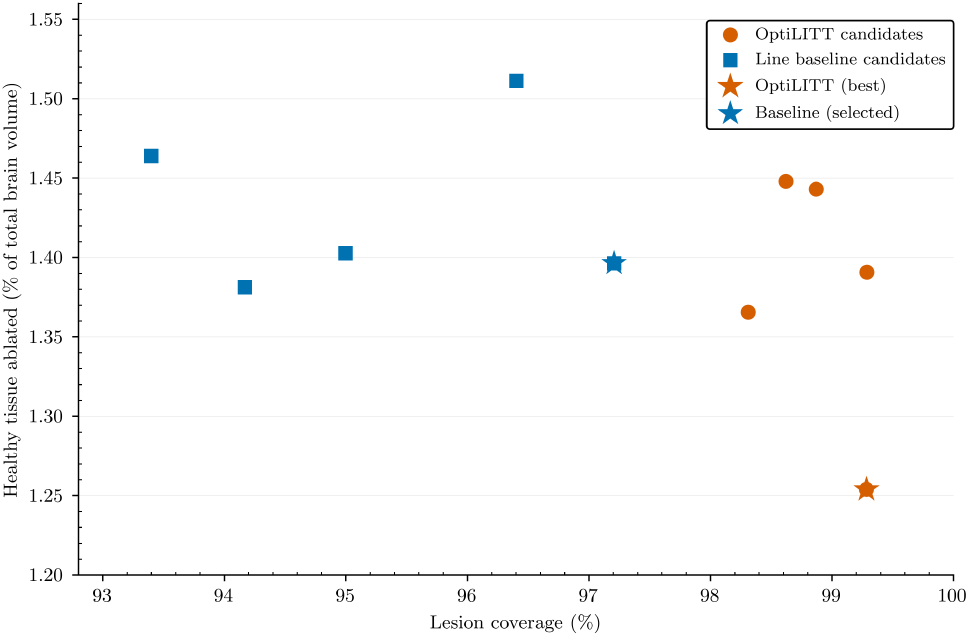
Coverage–healthy-tissue trade-off across the top five candidates per method.

### 4.2 Algorithmic Benchmarking

We benchmark our differential-evolution (DE) optimizer against four established search strategies on one hippocampal objective and parameter space (Section 3.3). The baselines comprise CMA-ES, Nelder–Mead and Powell with random multistart, and uniform random sampling as a control. All five methods minimize *OptiLITT* ‘s composite objective over an identical budget of 12,000 objective evaluations, replicated across 20 random seeds.

DE attains the lowest median final objective (117.4) with the tightest distribution across random seeds, outperforming CMA-ES (119.8), Powell (123.4), random search (123.5), and Nelder–Mead (125.4) (Figure 6d). DE’s advantage over each of the four baselines is statistically significant under a Holm-corrected Mann–Whitney *U* test (*p*_Holm_ ≤3.5 × 10^−^6). The pairwise win-rate matrix (Figure 6f) further reinforces this ranking, with a DE run outperforming a CMA-ES run selected at random with probability 0.93, and outperforming each of the remaining baselines with probabilities ranging from 0.99 to 1.00. This advantage becomes increasingly pronounced at higher target thresholds. Targets are defined as fractions of the composite-objective improvement from a trivial reference point (the median objective of a single sampled candidate) to the best objective value attained by any method. At the 99% target threshold, DE succeeds in 17*/*20 seeds, compared with 2*/*20 for CMA-ES and 0*/*20 for Powell, Nelder–Mead, and random search within the same evaluation budget (Figure 6b). This advantage, however, does not manifest at early evaluation budgets. At 100 evaluations DE (143.7) and CMA-ES (144.0) obtain comparable objectives, with Powell leading at 136.4 through 1,000 evaluations, and DE overtaking Powell only at 3,000 evals. From 3,000 to 12,000 evaluations, DE reduces its median composite objective by 6.6 units, outpacing both CMA-ES (5.7) and Powell (1.9), consistent with local search stagnating at a single optimum while DE’s mutation and crossover sustain exploration across the full solution space.

**Figure 6:**
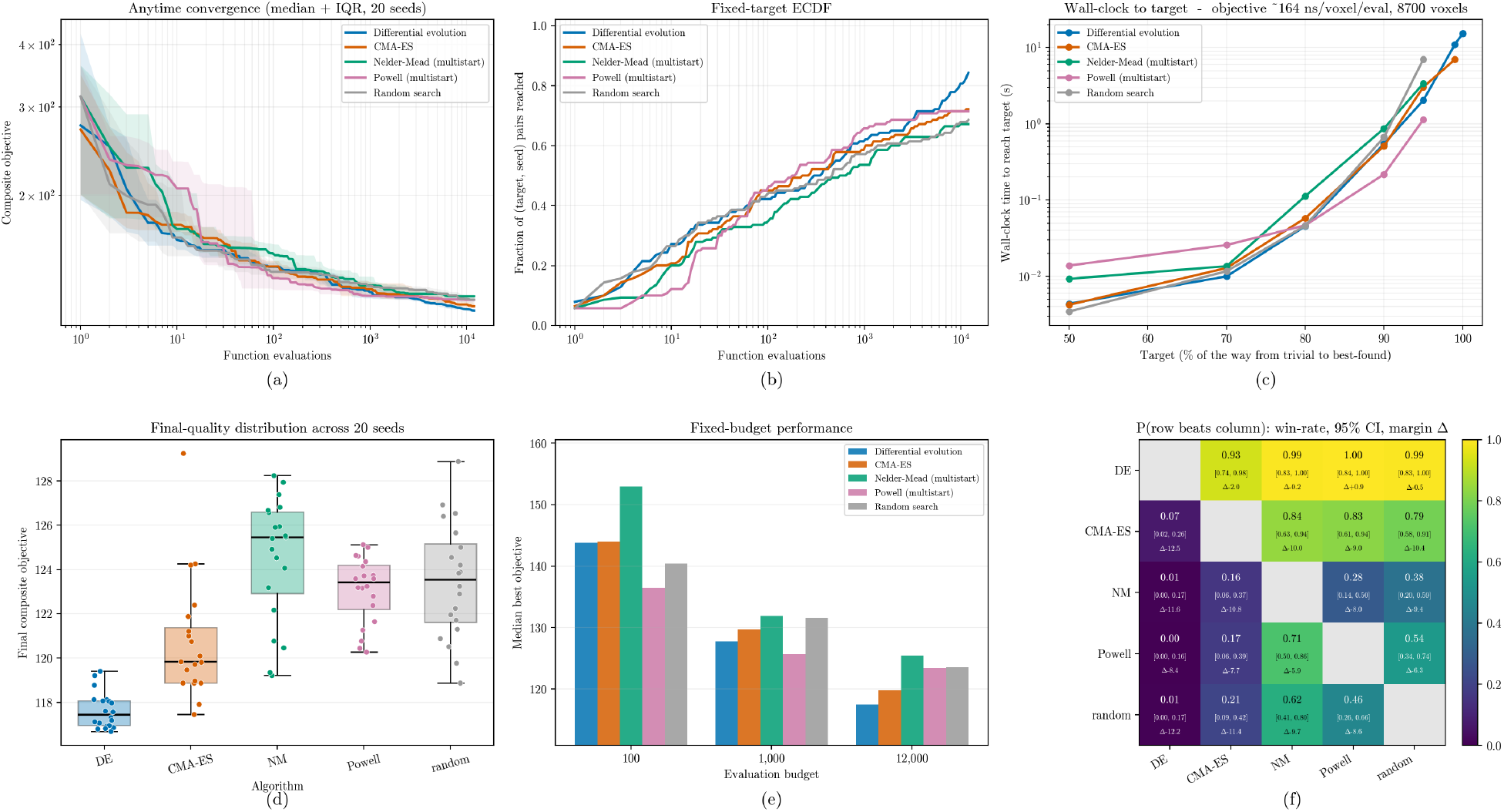
Benchmarking of five gradient-free optimizers on the composite trajectory objective (∼164 ns/voxel/eval, 8,700 voxels, 20 random seeds). (a) Anytime convergence: median composite objective vs. function evaluations with IQR bands. (b) Fixed-target ECDF: fraction of (target, seed) pairs for which the target was achieved vs. evaluation budget. (c) Wall-clock time to reach a target threshold, defined as a percentage of the objective improvement from the trivial baseline to the best solution found. (d) Final composite objective distribution across 20 seeds. (e) Fixed-budget median best objective at 100, 1,000, and 12,000 evaluations. (f) Pairwise win-rate matrix of outperformance probabilities between optimizers.

### 4.3 Limitations and Future Directions

#### Imaging scope and automation

The current pipeline accepts only axial and sagittal T1 DICOM stacks, and critical avoidance structures such as vasculature, ventricles, and white-matter tracts must be accounted for manually by the user. Incorporating complementary modalities such as T2/FLAIR, DTI tractography, and contrast-enhanced sequences may enable automated segmentation of these structures, allowing avoidance constraints to be enforced directly within the optimization.

#### Cohort generalization

Validation is currently limited to mesial-temporal (hippocampal) targets and a single retrospective comparison against an expert-planned trajectory. Broader claims require expansion across additional epileptogenic locations and all three lesion classification types. However, cohort growth is contingent on patient recruitment and procedural timelines, making large-scale multi-patient validation inherently slow to obtain.

#### Directional thermal modeling

`OptiLITT` models each ablation zone as a radially symmetric cylinder. This matches diffusing-tip applicators, but not directional side-firing probes that deposit energy to one side and can be rotated intraoperatively to conform to non-globoid lesions [12]. Extending the ablation primitive from a full cylinder to an angularly-restricted sector, parameterized by firing direction and arc width, would allow `OptiLITT` to plan for such probes by adding a per-fiber orientation parameter, leaving the coverage and spillover formulation otherwise unchanged.

#### Pediatric applicability

*OptiLITT* is currently intended for adult patients. Extension to pediatric populations is a planned future direction, though it will require higher geometric precision given smaller anatomy and tighter tolerances, and may necessitate re-tuning of optimization parameters such as fiber radii and entry-region constraints.

## Data Availability

All data produced in the present work are contained in the manuscript.

## Acknowledgements

We thank Sarah Johnson and Jessica Weber for their clinical assistance, and Stephan Bickel for his mentorship and resources.

## Data Availability Statement

The `OptiLITT` source code is not publicly available pending intellectual property disclosure. Anonymized patient imaging data and findings supporting the results of this study are otherwise contained within the manuscript.

## Author Contributions

**Natalie Yeung:** Methodology, Software, Validation, Formal analysis, Data curation, Writing – original draft, Visualization; **Akash Mishra:** Conceptualization, Data curation, Writing – original draft, Writing – review & editing; **Ashesh Mehta:** Conceptualization, Supervision, Writing – review & editing.

## Ethics Statement

This study involved a retrospective MRI analysis that carried no risk to participants. The study was approved by the Institutional Review Board at the Feinstein Institutes for Medical Research at Northwell Health and conducted in accordance with the ethical principles of the Declaration of Helsinki. Informed consent was obtained prior to participation.

